# “Antimicrobial Stewardship Knowledge Gaps among Healthcare Professionals at a Ghanaian Tertiary Hospital: a Cross-Sectional Study”

**DOI:** 10.64898/2026.05.04.26352400

**Authors:** Barbara Kobi Kyei, Emmanuel Boateng Kyei, Moses Yeboah Addo, Etepe Dugah, Charlotte A. T. Adu, Augustine Yeboah, Anne Boakyewaa Anokye Kumatia

## Abstract

The inappropriate use of antimicrobials enhances antimicrobial resistance (AMR). Antimicrobial stewardship (AMS) is a coordinated effort of prescribers, pharmacists, and nurses. Still, local data regarding AMS-related knowledge, attitudes, and practices (KAP) are scarce in many low and middle-income countries. We evaluated KAP regarding AMS among the healthcare providers at Komfo Anokye Teaching Hospital (KATH), Ghana, and found the related factors. A cross-sectional survey in the form of a descriptive survey was conducted among medical doctors, pharmacists, and nurses at KATH. Knowledge, attitude, and practice were evaluated using a structured questionnaire. The scores were converted into percentages and classified as good (>=60%) or poor (<60%). Chi-square tests were used to test associations, and logistic regression to predict good KAP (p<0.05). A total of 349 healthcare professionals participated, which comprised: 91 medical doctors (26.1%), 101 pharmacists (28.9%), and 157 nurses (45.0%). The majority of the respondents had formal AMS/AMR training (69.6%), and 37.0% had updated training the previous year. Only 18.6% demonstrated good AMS-related knowledge, although attitudes were largely positive (95.7% good) and reported practices were mostly appropriate (77.4% good). In multivariable models, greater years of practice (5–9 years: adjusted odds ratio [AOR] 2.32; >=15 years: AOR 2.77) and formal training (AOR 2.94) were associated with good knowledge. Formal training was also associated with good attitudes (AOR 5.19). Compared with medical doctors, nurses had lower odds of good practice (AOR 0.29), while pharmacists had higher odds (AOR 1.41). Participants with 10–14 years of experience had higher odds of good practice (AOR 3.18). This study revealed that marked knowledge deficits exist, despite favourable attitudes and generally good self-reported AMS practices. Role-tailored, competency-based AMS training with regular updates and reinforcement through practical stewardship tools is needed to translate positive attitudes into evidence-based prescribing and administration behaviours.

## Introduction

Antimicrobial resistance (AMR) is a significant global epidemic that undermines the capacity of health systems to prevent and treat infections [1–3]. Resistant infections are also linked to length of illness, increased complications and death rates, and escalated health care expenses. The impact of bacterial AMR worldwide is huge, and it is even worse in low- and middle-income nations, where the incidence of infectious diseases is high, and health systems in these countries are usually limited by laboratory capacity, the availability of quality medicines, and infection prevention resources [2]. These issues may increase the choice and transmission of resistant organisms in hospital environments and, as such, AMR serves as a patient safety concern and a health system performance problem. One of the causes of AMR is the improper use of antimicrobials. The selection pressure is also heightened when antimicrobials are initiated without an apparent indication, selected in a broad spectrum, administered in inappropriate doses or durations, or the antimicrobial is administered without following treatment recommendations [1, 4, 5]. Diagnostic uncertainty, inaccessibility to timely microbiology findings, shortages of essential medicines, and differences in clinical training and supervision also constitute additional factors affecting prescribing decisions in most hospitals. These facts complicate the ability to have a consistent level of rational use of antimicrobials in the absence of a deliberate system and workforce intervention.

Antimicrobial stewardship (AMS) offers a viable model of enhancing antimicrobial use by integrating concerted efforts to aid in the best choice, dosing, route, and treatment duration [6, 7]. In various settings, there is evidence that effective stewardship has the potential to enhance the quality of care, decrease the unnecessary exposure to antimicrobials, decrease the incidence of some resistant infections, and enable improved clinical outcomes [8].

Stewardship is most effective when it is multidisciplinary and embedded within routine clinical workflows, combining education and guidelines with tools such as audit and feedback, review of therapy after initiation, and access to local resistance patterns to guide empiric choices.

In Ghana, AMR has been reported across significant bacterial pathogens, and AMR and the related national policy contexts, such as the National Action Plan on AMR, highlight enhanced stewardship, as well as infection prevention and control [5, 9]. The translation of national priorities into the tangible improvements of the hospital level requires an informed and engaged healthcare workforce. Medical doctors, pharmacists, and nurses impact antimicrobial decisions at various stages of the care pathway, whether it is initiating treatment, dispensing, administering, monitoring, or reinforcing compliance with guidance. For this reason, understanding knowledge, attitudes, and everyday practices related to AMS within these professional groups is essential for designing interventions that are realistic, role-specific, and likely to be sustained.

This study assessed AMS-related knowledge, attitudes, and practices among medical doctors, pharmacists, and nurses at Komfo Anokye Teaching Hospital (KATH), Ghana, and examined factors associated with good performance.

The findings are intended to identify priority gaps for targeted training and support, and to inform the design of stewardship strategies that strengthen rational antimicrobial use within a large tertiary referral hospital setting.

## Methods

### Study design and setting

A descriptive cross-sectional study was conducted at KATH in Kumasi, Ghana. The study targeted professional healthcare groups that are directly involved in antimicrobial prescribing, dispensing, and administration.

### Study population and sample size

The study population comprised medical doctors, nurses, and pharmacists working at KATH who consented to participate. The sample size was estimated using Cochran’s formula for a single proportion with 95% confidence level, 5% margin of error, and an assumed prevalence of 50%, yielding an initial sample of 384. A finite population correction was applied, resulting in a target sample size of 350. A total of 349 responses were analysed, as one participant opted out.

### Data collection

Data were collected using a structured questionnaire adapted from previously published AMS/ AMR KAP tools [10, 11]. The instrument comprised four sections: socio-demographics, knowledge of AMS interventions, attitudes towards AMS, and AMS-related practices. Before administration, a pre-test was conducted at a different facility, and the feedback informed minor revisions.

### Outcome definitions and scoring

Knowledge was scored as 1 for a correct response, and 0 for an incorrect or ‘Not sure’ response (maximum 9 points). Attitude items were scored from 5 (strongly agree) to 1 (strongly disagree) for positively worded statements, with reverse scoring for negatively worded statements. Practice items were scored as 1 for appropriate practices and 0 for otherwise (maximum 5 points).

For each domain, the score was converted to a percentage and categorized as good ( ≥ 60%) or poor (< 60%).

### Statistical analysis

Data were entered in Microsoft Excel and analysed using R. Descriptive statistics were summarized as frequencies and percentages. Associations between sociodemographic variables and KAP categories were assessed using chi-square tests. Variables were entered into logistic regression models to estimate crude odds ratios (OR) and adjusted odds ratios (AOR) with 955 confidence intervals (CI) for predictors of good knowledge, attitude, and practice. Statistical significance was set at p < 0.05.

### Ethics statement

Ethical approval was obtained from the Komfo Anokye Teaching Hospital Institutional Review Board (KATH IRB/AP/142/24). Additionally, administrative approval was obtained from the management of the Komfo Anokye Teaching Hospital to ensure institutional support. Participation was voluntary, written informed consent was obtained from all participants, and responses were kept confidential.

## Results

### Participant characteristics

Out of 349 respondents, 203 (58.2%) were females, and 146 (41.8%) were males. Under professional distribution, there were 91 medical doctors (26.1%), 157 nurses (45.0%), and 101 pharmacists (28.9%). Most participants had 0–4 years of practice (43.6%), while 10.9% had >=15 years of experience. Formal training on AMR/AMS was reported by 243 participants (69.6%), and 129 (37.0%) reported update training in the last year (Table 1).

**Table 1.** Socio-demographic characteristics of participants (N=349)

| Variable(n=349) | Frequency | Percentage (%) |
| --- | --- | --- |
| **Sex** |  |  |
| Male | 146 | 41.83 |
| Female | 203 | 58.17 |
| **Professional Designation** |  |  |
| Medical Doctor | 91 | 26.07 |
| Nurse | 157 | 44.99 |
| Pharmacist | 101 | 28.94 |
| **Years of Practice** |  |  |
| 0 -4 | 152 | 43.55 |
| 5-9 | 84 | 24.07 |
| 10-14 | 75 | 21.49 |
| $\geq 15$ | 38 | 10.89 |
| **Formal Training** |  |  |
| Yes | 243 | 69.63 |
| No | 106 | 30.37 |
| **Update Training** |  |  |
| Yes | 129 | 36.96 |
| No | 220 | 63.04 |

### Assessment of Knowledge, attitudes and practices

In all, 65 (18.6%) of participants had a good knowledge score, as 284 (81.4) had poor knowledge. Contrastingly, attitudes towards AMS were positive, with 334 (95.7%) of participants categorized as having a good attitude. Most of the participants, 270 (77.4%), also reported good AMS-related practices. (Table 2).

**Table 2.** Levels of AMS-related knowledge, attitude, and practice (N=349)

| Domain | Good, n (%) | Poor, n (%) | Cut-off |
| --- | --- | --- | --- |
| Knowledge | 65 (18.6) | 284 (81.4) | $\geq 60\%$ |
| Attitude | 334 (95.7) | 15 (4.3) | $\geq 60\%$ |
| Practice | 270 (77.4) | 79 (22.6) | $\geq 60\%$ |

### Bivariate associations

Professional designation was significantly associated with knowledge (p=0.018), attitude (p=0.032), and practice (p=0.002). Years of practice was associated with attitude (p=0.009) and practice (p=0.033), but showed a borderline association with knowledge (p=0.059). Former training on AMR/AMS was associated with better knowledge (p=0.002), and attitude (0.005), but not practice (0.486) (Table 3a-3c).

**Table 3.**
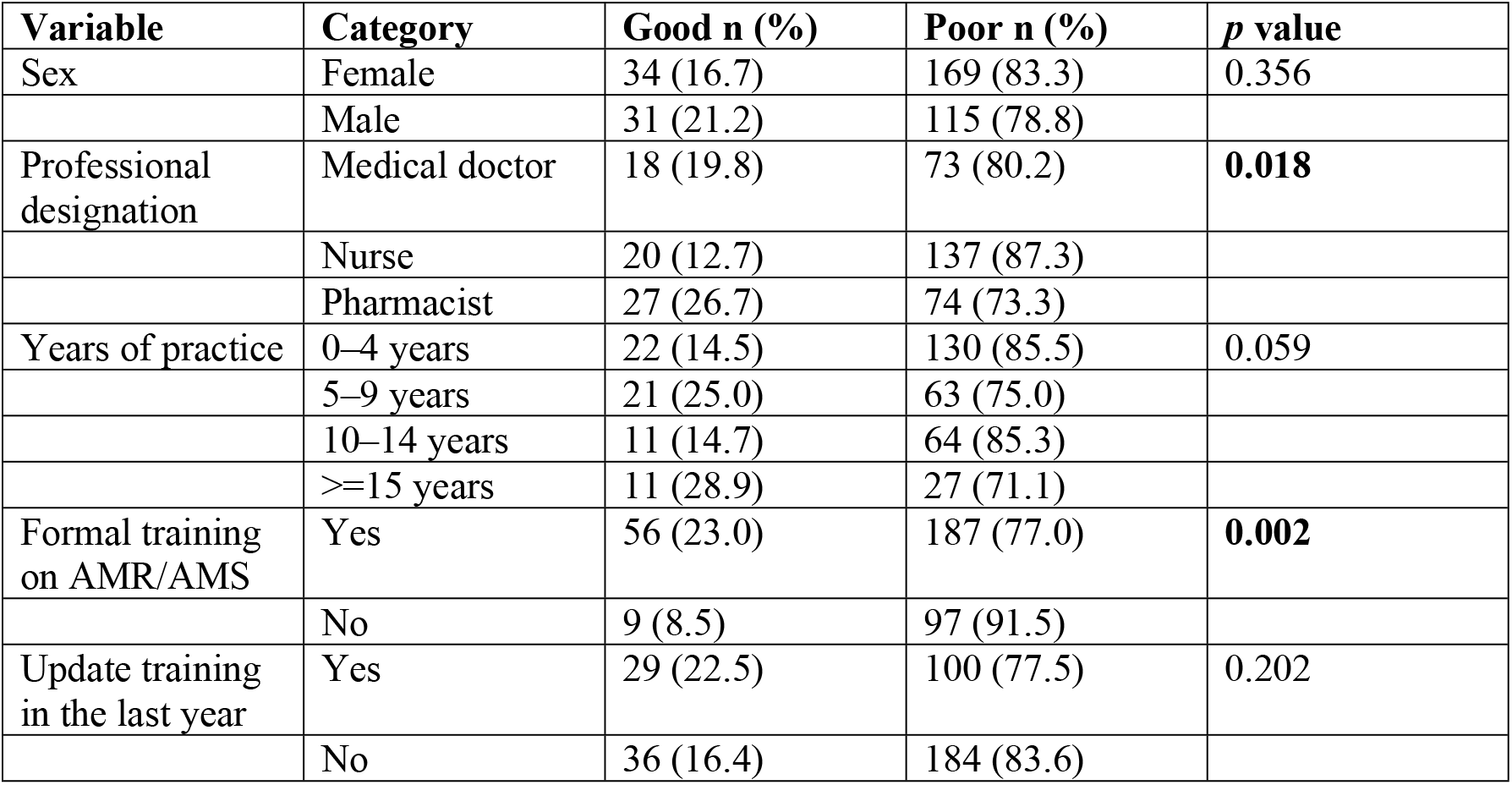
Factors associated with knowledge level.

**Table 4.**
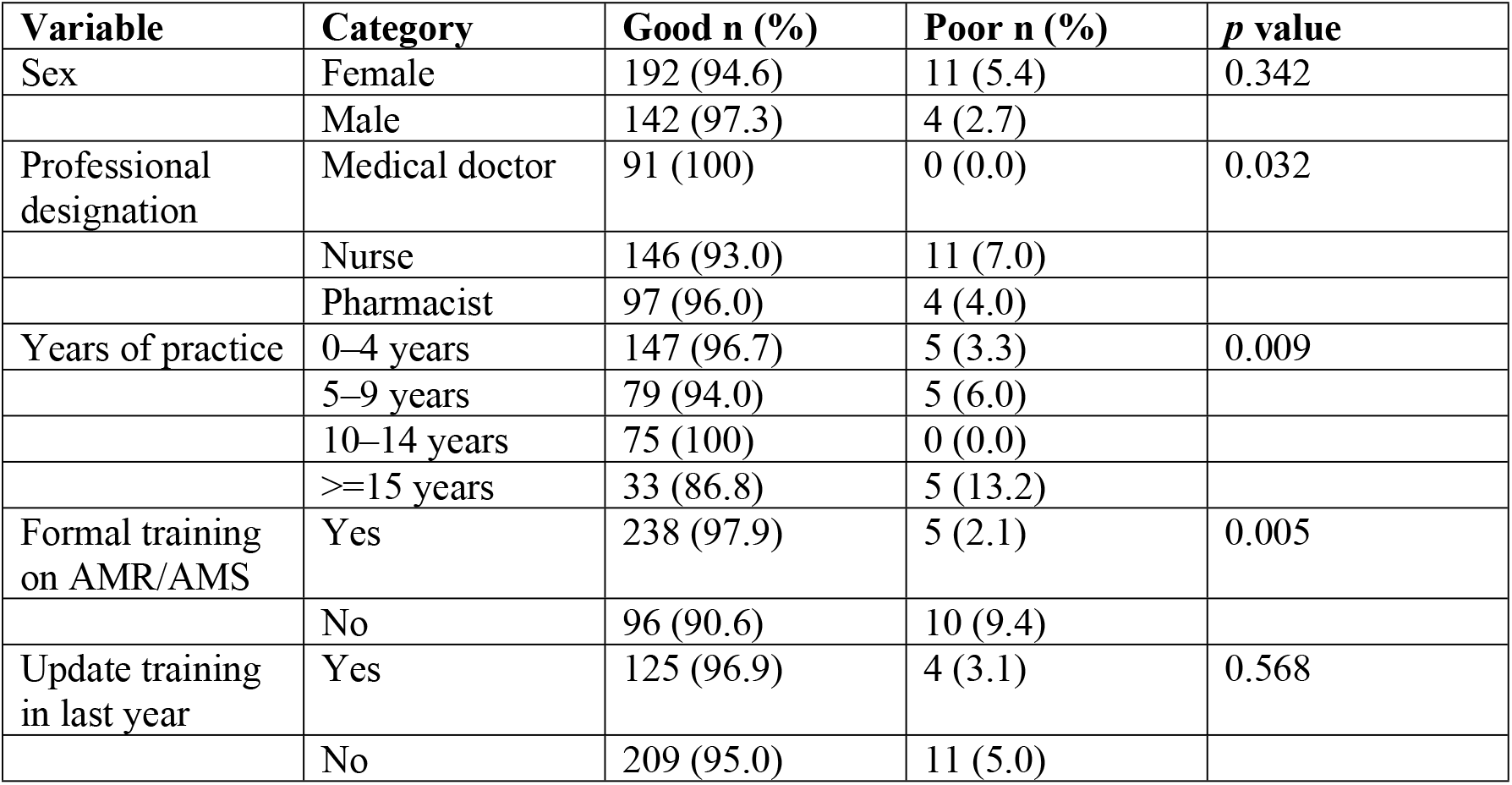
Factors associated with attitude level.

**Table 5.**
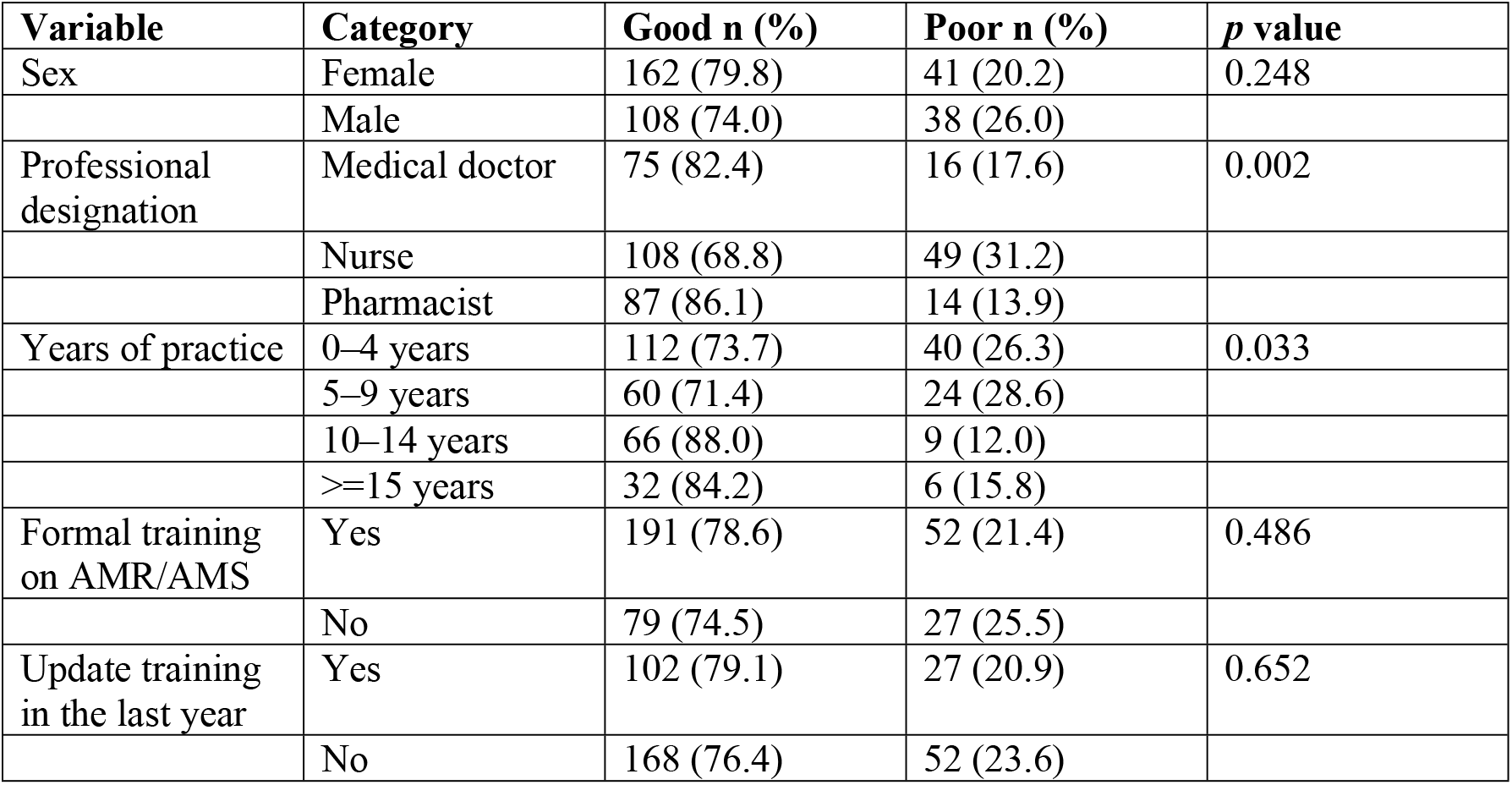
Factors associated with practice level.

### Multivariable predictors of good KAP

Knowledge-wise, participants with 5-9 years of experience (AOR 2.32; p = 0.023), those with 15 or more years of experience (AOR = 2.77; p = 0.022), and those who had had formal training (AOR = 2.94; p = 0.013) were independently associated with good knowledge. In terms of attitudes, only formal training was found to be statistically significant (AOR = 5.19; p = 0.033). In terms of practice, nurses had significantly lower odds of good practice (AOR: 0.29; p = 0.001) compared to medical doctors, while pharmacists showed high odds (AOR: 1.41; p = 0.006). Additionally, participants with 10 to 14 years of professional experience had high odds of good practice (AOR=3.18; p = 0.006) (Table 6).

**Table 6.**
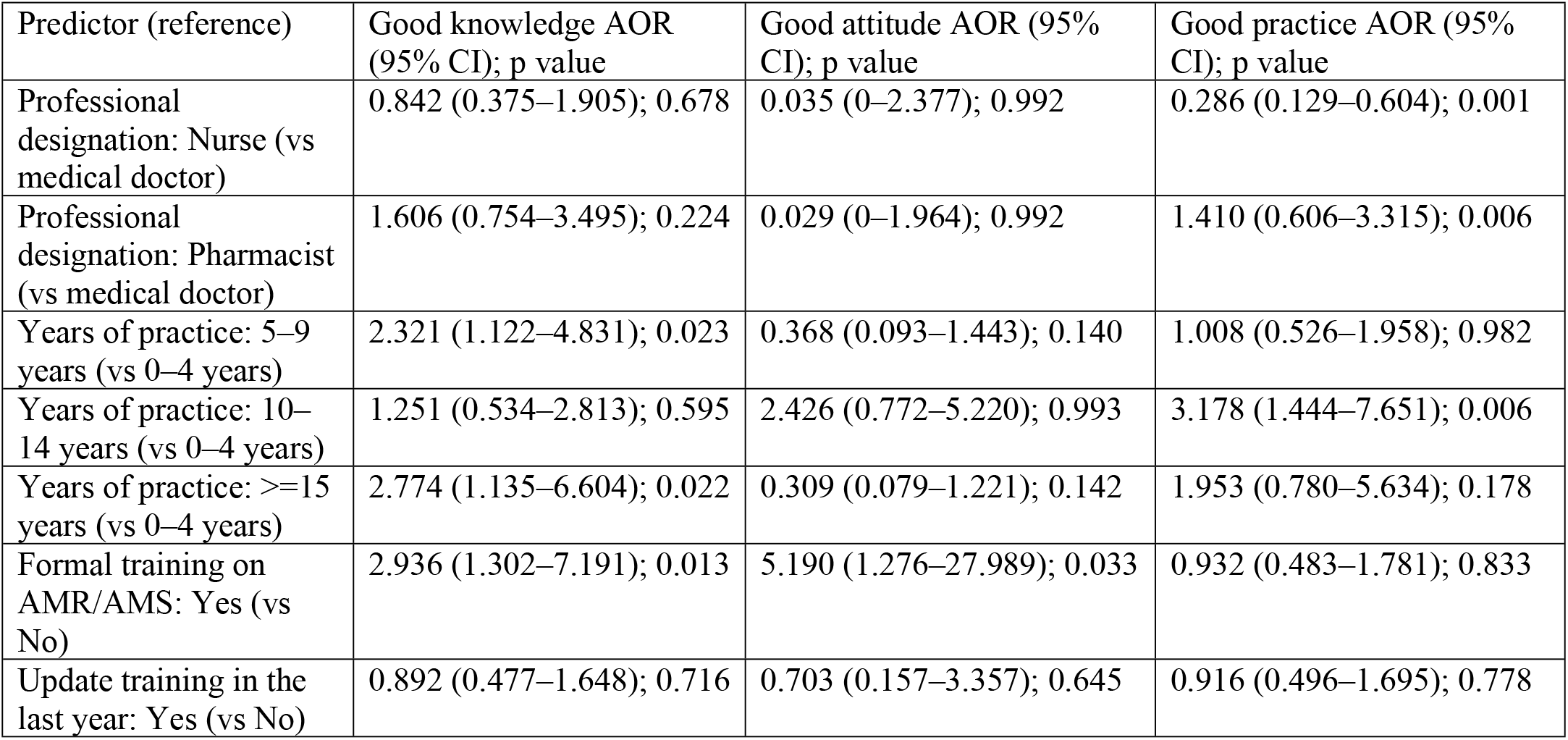
Adjusted odds ratios (AOR) for predictors of good knowledge, attitude, and practice.

## Discussion

This study demonstrates a gap in knowledge, attitude, and practice among healthcare professionals. Knowledge was low, while attitudes were high, and self-reported practices were generally good. This pattern suggests a strong recognition of antimicrobial stewardship as a priority among professionals, but practical stewardship tools may not be understood and consistently integrated into everyday clinical decision-making.

Low knowledge has also been reported across hospital settings in Ghana and other African countries, although estimates vary by tool, cadre-mix, and scoring thresholds. A recent multicentre survey in Ghana reported even lower proportions of “good” knowledge, with moderate practice levels, despite generally positive attitudes [12]. Similarly, in a Ghanaian tertiary hospital, healthcare providers described limited familiarity with AMS strategies and guideline-based decision-making before targeted educational activities [13]. These findings indicate that knowledge deficits are common and may persist even when national AMR policies and institutional messaging have improved awareness of the threat of AMR.

The domain of knowledge evaluated by this study included practical stewardship processes such as prospective audit-and-feedback, the use of antibiograms, prior authorisation, / stop orders, and point-prevalence surveys, and current global stewardship in use, like the WHO AWaRe framework and Access antibiotic targets. These are not theoretical but ‘working bits’ of stewardship programmes allowing prescribers and teams to choose narrower agents when it is suitable, where to review, de-escalate under the results, and review progress. Hence, the observed knowledge gap in this study could be due to inadequate exposure to organised stewardship processes, insufficient access to local decision support resources, and inconsistent diagnostic support-barriers, which repeatedly emerge in Low- and Middle-Income Countries (LMIC) stewardship studies [14].

In our cohort, we found that formal education in AMR/AMS and increased professional experience were associated with higher odds of good knowledge and attitudes. This is consistent with the intervention evidence from Ghana, indicating that targeted stewardship training can enhance confidence and perceived competence in AMS roles in clinicians and other professionals [15]. However, “updated training in the last year” was not independently associated with KAP outcomes. This could be due to heterogeneity in training content, duration, and reinforcement; brief, didactic sessions. This may increase awareness without meaningfully building applied competence.

Recent studies show that AMR stewardship education is most effective when it is ongoing, case-based, linked to local guidance and feedback, and embedded within a wider package of enabling interventions [14, 16, 17].

Positive attitudes were high in this current study, and align with other surveys of African hospitals where healthcare workers usually support AMR stewardship objectives despite variable knowledge and practice [12, 18, 19]. However, high attitude scores do not translate into behaviour when stewardship roles in multidisciplinary groups are not well defined, as widely reported in LMIC settings [14, 20].

Professional role differences were prominent for practice. Nurses had lower odds of good stewardship practice compared with medical doctors, while pharmacists had higher odds. This aligns with evidence that nursing engagement in AMS is often under-recognized and constrained by role ambiguity, hierarchical decision-making, and limited access to prescribing discussions [21, 22].

The high reported practice among pharmacists in this study supports the evidence of pharmacist-led stewardship activities in sub-Saharan Africa [23]. A systematic review of pharmacist-led AMS programs in sub-Saharan Africa concluded that pharmacists can deliver improvements in antimicrobial use and stewardship indicators [23].

From an implementation standpoint, findings from this study point to a need for stewardship strategies that go beyond awareness-raising. The WHO practical toolkit for AMS in healthcare facilities emphasises a stepwise approach: establish governance and a multidisciplinary team, implement context-appropriate interventions, and build monitoring systems [24].

## Strengths and limitations

This study is one of the few studies to include multiple professional groups that are central to AMR stewardship, while involving a structured and widely accepted instrument with defined scoring criteria. However, the limitation of this study was the reliance on self-reported practices, which could be affected by social desirability bias.

## Conclusion

This KAP analysis revealed that healthcare professionals demonstrated very positive attitudes and self-reported good AMS practices, yet a substantial knowledge gap persists. Formal AMS/AMR training and experience were associated with improved knowledge and attitudes, while professional designation influenced practice. Targeted and role-specific AMR stewardship training with reinforcement through facility tools and multidisciplinary structures is recommended.

## Declarations

### Ethical approval and consent to participate

Ethical approval was obtained through the Komfo Anokye Teaching Hospital Institutional Review Board (KATH IRB/AP/142/24), with approval from Hospital management. Written informed consent was obtained from all participants.

### Data Availability

The dataset and questionnaire may be made available from the corresponding author on request, subject to institutional approvals.

### Author’s contributions

BKK conceived the study, developed the protocol, and the data collection tool. EKB contributed to data cleaning and interpretation of findings. MYA supported the statistical analysis plan and interpretation of results and contributed to the revision of the manuscript. CATA and AY performed data management and statistical analysis, interpreted findings, and drafted the initial manuscript. ABAK and ED provided methodological guidance and supervision throughout the study and reviewed the manuscript. All authors contributed to manuscript revision, read and approved the final version, and agree to be accountable for all aspects of the work.

## Acknowledgements

The authors thank the staff of Komfo Anokye Teaching Hospital who participated in the survey, and the hospital management for their support.

## List of abbreviations

AMR: Antimicrobial resistance
AMS: Antimicrobial stewardship
AOR: Adjusted odds ratio
CI: Confidence interval
HCP: Healthcare professional
KAP: Knowledge, attitudes, and practices
KATH: Komfo Anokye Teaching Hospital
LMIC: Low- and Middle-Income Countries
OR: Odds ratio
WHO: World Health Organization

